# Effects of robotic gait rehabilitation with Lokomat ® in vascular hemiplegia: a single-center retrospective observational study

**DOI:** 10.1101/2024.07.12.24310354

**Authors:** Constance Michel-Sayeg, Soizic Injeyan, Xavier Ropero, Olivier Arkoun, Rémy De Mil, Anne Peskine

## Abstract

**Introduction:** Gait rehabilitation after a stroke is a major concern in physical medicine and rehabilitation. Since 2007, « Le Normandy» rehabilitation center is equipped with robotic assisted gait training named “Lokomat”; vascular hemiplegic patients benefit from it. This study was conducted to investigate the effect of Lokomat on the walking performance of hemiplegic patients after a stroke and to identify the influence of certain factors.

**Methods:** This was a retrospective, observational study conducted at « Le Normandy » rehabilitation center in Granville, France. All adult patients hospitalized between 2007 and 2018 for a first stroke within the last six months who completed a full Lokomat program were included. Medical data were collected : sex, age, type of stroke, time since the onset, modified Functional Ambulation Category (mFAC), date of stroke and results of the six minutes walking test (6MWT) before and after rehabilitation. The outcome was the difference between the 6MWT before and after the rehabilitation to determine the evolution of the walking capacity and factors associated with a gain.

**Results:** We included 235 patients. The median gain on the 6MWT was 18 meters (p<0,01). Two factors were significantly associated with the 6MWT gain : rehabilitation phase and severity of stroke. The median gain was 21.22 meters (95% CI [1.86 ; 44.93]) for patients in early rehabilitation phase versus late rehabilitation phase. The median gain was 34.22 meters (95% CI [16.20 ; 74.69]) for ambulatory patients at the onset compared to non-ambulatory patient. There were no significant difference of median gain between men and women (3.22 meters 95% CI [−11.86 ; 19.48]), between ischemic and hemorrhagic strokes (1.67 meters 95% CI [−10.59 ; 17.03]) and between the post-2016 and pre-2016 periods (6.22 meters 95% CI [−7.99 ; 32.78]). There was a decrease of 0.56 meters (95% CI [−0.99 ; 0.16]) per year at the 6MWT, and this result was not significant.

**Conclusion:** There was a significant improvement in the 6MWT after robotic rehabilitation by Lokomat. This improvement was greater in patients in early rehabilitation phase and when the patient was initially ambulatory.

## Introduction

A stroke is a sudden loss of neurological function due to an ischemic or hemorrhagic intracranial vascular event (1). Each year, an estimated 140 000 patients in France are hospitalized for a stroke, which is the second cause of death and the first cause of long-term disability in adults (2). At one year, it is estimated that 20% of patients die. Walking dysfunction occurs in more than 75% of survivors, and 25% do not recover at 3 months (3).

Walking is an important indicator of the activities and participation domains of the International Classification of Functioning, Disability, and Health (ICF) (4). Regaining the ability to walk is a very important goal for patients after a stroke.

Rehabilitation is a complex process requiring multidisciplinary care, including the patient and their goals, family and friends, other caregivers, physicians, nurses, physical and occupational therapists, speech-language pathologists, recreation therapists, psychologists, nutritionists, social workers, and others (5).

Robotic-assisted gait training (RAGT) has been used since 1980 to assist patients with movement dysfunction caused by neurological disorders (6). Electromechanical-assisted gait training and RAGT can be used in walking rehabilitation (7) and provide repetitive task training (RTT). Electromechanical devices can be differentiated into end-effectors and exoskeleton devices. Examples of end-effectors are “G-EO System,” “Lokohelp,” and “Gait Trainer.” Examples of exoskeletons are “LOPES” (lower extremity powered exoskeleton) and “Lokomat” (8). Several reviews have shown that people who receive RAGT in combination with physiotherapy after a stroke are more likely to achieve independent walking than people who receive gait training without these devices but are not likely to improve their walking speed or walking capacity. However, the type of RAGT and the analyzed population were not clear (10) (11).

Since 2007, the rehabilitation center of Granville Le Normandy has been equipped with a Lokomat (Hocoma, Volketswil, Switzerland), which is a robotic-assisted gait trainer with treadmill training and body weight support. The main difference with treadmill training is that the patient’s legs are guided by a robotic device controlled by a computer.

Previous research, carried out particularly on the Lokomat, has found mixed results for gait recovery and has not found superior effects of robotic training compared to conventional therapy on walking outcomes (12), (13) but has shown that Lokomat could be safely used at no detriment to patient outcomes (14).

The objective of this observational study was to assess the effect of Lokomat® rehabilitation on the walking capacity of hemiplegic patients. The influence of certain factors was also studied.

## Methods

### Study design

This was a retrospective, observational study conducted at the Le Normandy Rehabilitation Center in Granville, France. Patients selected were hospitalized in the center for rehabilitation after an ischemic or hemorrhagic stroke between 2007 and 2018.

The inclusion criteria for this study were:

1. adult age 18 or older
2. first hemiparetic stroke based on clinical examination or magnetic resonance imaging (MRI)
3. occurrence of the stroke less than 6 months before rehabilitation
4. The patient should have trained with Lokomat for a first complete program (including 20 sessions); eventual subsequent sessions were not taken into account.

The exclusion criteria were:

1. recurrent stroke
2. non-complete session of Lokomat

All patients who had a Lokomat session were included in a Lokomat database stating their pathology. All vascular hemiplegic patients were eligible to use the Lokomat unless they had one of the following contraindications: obesity (more than 130 kg), major cognitive impairment, hemianopia, any co-morbidity or disability other than hemiparesis that would preclude gait training, a urinary catheter or gastrostomy, or major spasticity.

The training program consisted of 20 sessions of Lokomat, 30 minutes per session, five days a week (from Monday to Friday) for four consecutive weeks. The first session allowed us to get familiar with the robotic equipment and take measurements for the installation.

All patients had conventional gait training in addition to the Lokomat. (15)(5) A trained physical therapist supervised the treatment and adjusted treadmill speed, guidance, and body-weight support. Levels of body weight support and guidance progressively decreased, and treadmill speed progressively increased (16).

Each patient benefited from a clinical examination by a doctor and a physiotherapist on arrival before starting rehabilitation as part of their care. No additional examinations or functional tests were performed if they were not indicated as part of the rehabilitation. All patients have agreed to undergo the therapy.

The protocol was approved by the ethics committee of the University Hospital of Caen, France.

### Definitions and measurements

Medical data were collected retrospectively from medical records and included: sex, age, type of stroke (ischemic or hemorrhagic), time since the onset, modified Functional Ambulation Categories (mFAC) before rehabilitation, date of the stroke, and results of the six-minute walk test (6MWT) before and after rehabilitation.

The 6MWT is commonly used in people with stroke undergoing rehabilitation (17) to classify a patient into a degree of functional limitation. The patient is asked by his physiotherapist to walk as fast as possible for 6 minutes, with or without a walking aid. The reference value is calculated by a formula including gender, height, and weight. For an adult, the normal range is 407–535 meters (19) (20) (21).

The mFAC (22) (Annex 1) is a scale that evaluates the capacity of ambulation and the need for technical assistance. Classes 0 and 1 define a patient not able to walk, and classes 2 up to 8 define a walking patient with or without technical assistance.

**Annex 1:** modified Functional Ambulation Category.

|  |
| --- |
| **Class 0** : Can not walk or needs assistance from more than one person. |
| **Class 1** : Can walk with the permanent assistance of a person. |
| **Class 2** : Can walk with intermittent assistance of one person. |
| **Class 3** : Can walk with verbal support without physical contact. |
| **Class 4** : Can walk alone on flat surfaces, but stairs are impossible. |
| **Class 5** : Can walk alone on flat surfaces. Passage of stairs is possible with physical assistance of a third person. (physical contact or simple supervision) |
| **Class 6** : Can walk alone on flat surfaces. Stairs can be negotiated using a ramp or cane, without assistance and/or supervision from a third party. |
| **Class 7** : Can walk alone on flat surfaces. Stairs can be passed alone but abnormally (slower with limping), without help and/or supervision from someone, nor external support. |
| **Class 8** : Can walk independently on level surfaces and negotiates stairs normally without the use of a handrail or cane with normal stair climbing. |

The mFAC was evaluated before the rehabilitation and was therefore a measure of the initial severity of the stroke in terms of motricity.

### Statistical Analysis

Our outcome was the difference between the 6MWT before and after rehabilitation to determine the evolution of the walking capacity and factors associated with an improvement. The results of the mFAC scale were categorized into two groups: ambulatory patients with or without technical assistance (mFAC 2 to 8) and non-ambulatory patients (mFAC 0 and 1). Modified FAC class 0 means non-walking. We decided that it meant a 6MWT of 0 meters. For other mFAC classes, missing data from the 6MWT was excluded.

Time since stroke was categorized into two groups, described as early rehabilitation (0–3 months) and late rehabilitation (3-6 months) for the acute phase after stroke (23): The date of stroke was categorized into two periods: before and after 2016. In 2016, the thrombectomy protocol was generalized and standardized at Caen University Hospital for all of our region (24) (25). We made the hypothesis that more thrombectomies were performed after 2016, possibly leading to a better prognosis for gait rehabilitation.

Categorical data are presented as counts and percentages. For continuous data, normality was tested using the Kolmogorov-Smirnov test and assessment of histograms. Distributions of all continuous data were skewed and were therefore described with medians and interquartile ranges (IQR) and then compared with the use of the Mann-Whitney test or the Spearman correlation coefficient, as appropriate. Bivariate analyses were performed to compare, first, the 6MWT before and after rehabilitation using a paired Mann-Whitney test, and then to assess the association between the gain in 6MWT and the following characteristics: sex, age, type of stroke, rehabilitation phase, mFAC before rehabilitation, and period.

We then fitted a multivariable quantile regression model using the modified version of the Barrodale and Roberts algorithm, and confidence intervals for the estimated parameters were produced by inverting a rank test without assuming that the errors were independent and identically distributed. We chose to model the 50% quantile, i.e., the median, which allows us to obtain the adjusted augmentation of the median gain in 6WMT for each variable and a 95% confidence interval. A quantitative regression model was used instead of an ordinary least squares (OLS) model because OLS residuals were non-normal, highly skewed, and heteroskedastic. The final multivariable model was based on all variables associated with the evolution of the 6MWT in bivariate analysis. The P-value was 0.20 for inclusion in the model. Age and sex were forced into the model regardless of their p-value as well as variables of clinical interest, including the type of stroke and the date of the stroke as a proxy of thrombectomy treatment.

All analyses were two-sided with a 5% type I error and were completed in R 4.1.3 software, using the quantreg » package for the quantile regression model.

## Results

We retrospectively reviewed 268 medical records with strokes in the Lokomat Database between 2007 and 2018.

Thirty-three records were excluded because there were no first strokes or first Lokomat sessions. Patients with incomplete Lokomat sessions were likewise excluded.

A total of 235 patients were included. When the results of the 6MWT before or after rehabilitation were missing, we could not calculate the difference. It was the case for 72 patients, and these patients was not included in the analysis (Figure 1).

**Figure 1:**
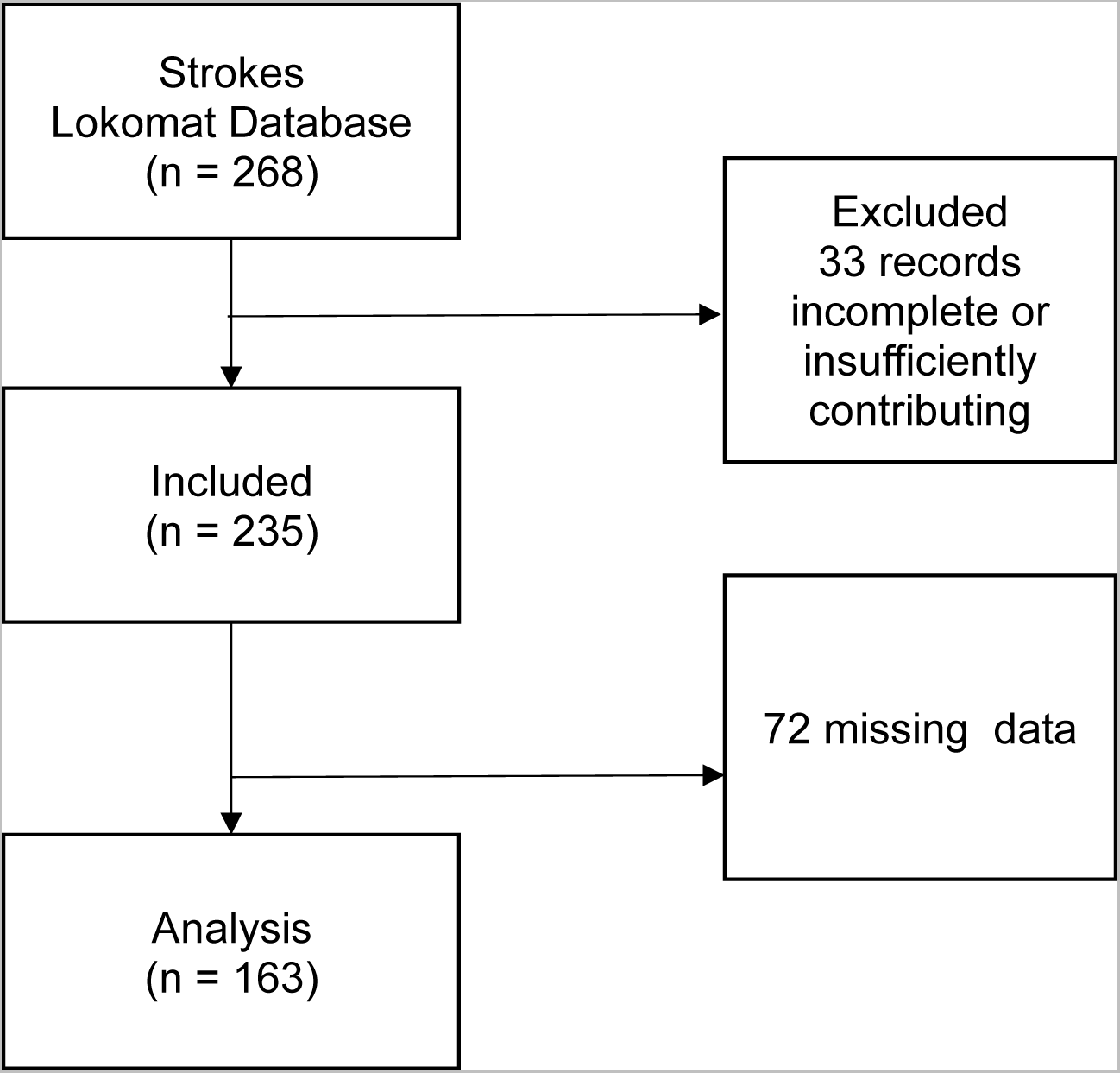
The flow diagram of the study.

**Figure 2:**
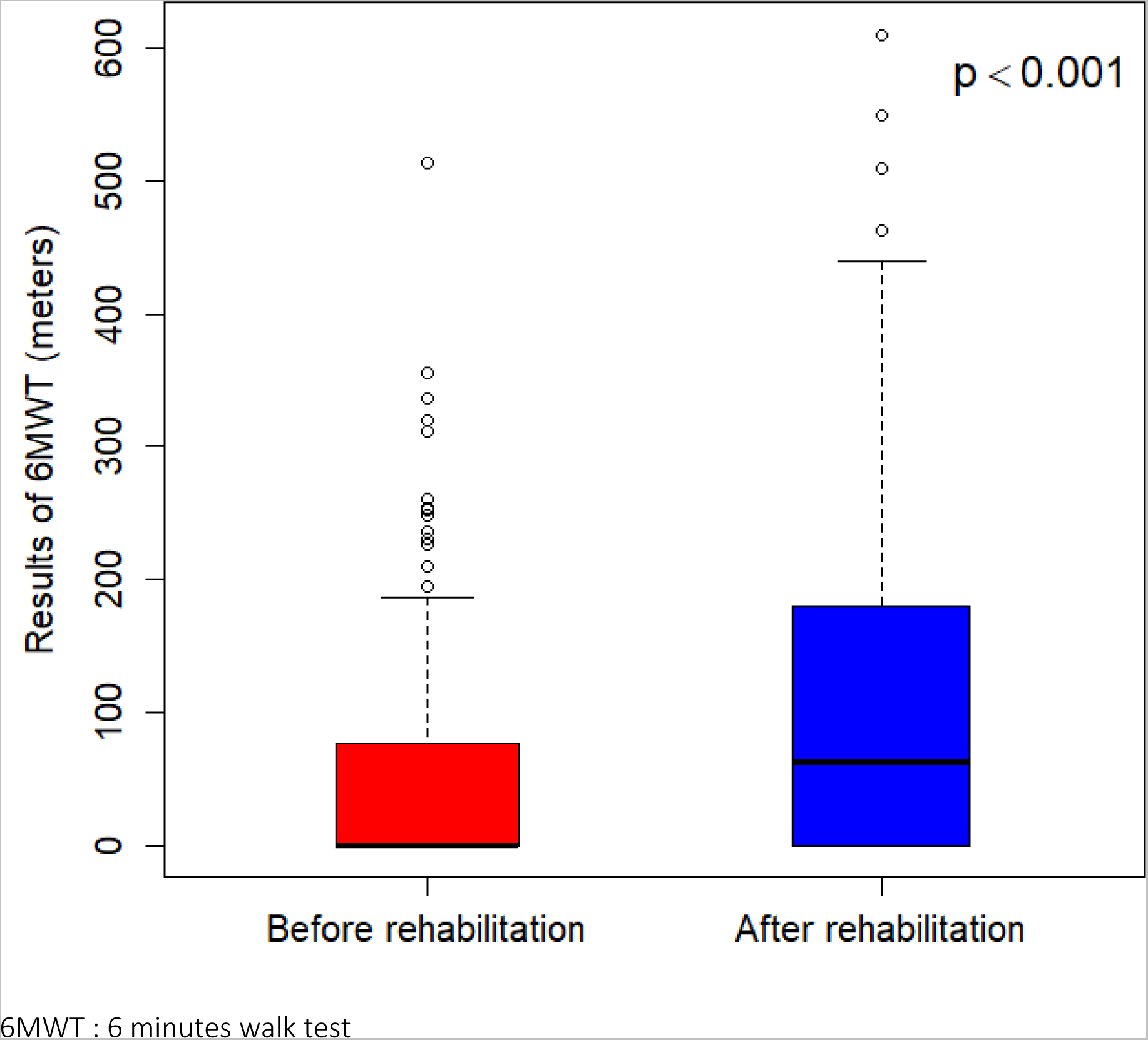
Results of the walking test.

There were 151 men, with a median age of 66 years. Sixty-eight percent had an ischemic stroke, and 73% of patients were in the early rehabilitation phase. There were 71% of non-ambulatory patients. The characteristics of the population are summarized in Table 1.

**Table 1:** Baseline characteristics of the patients.

| Variable | Value |
| --- | --- |
| **Subjects - n** | 235 |
| **Sex - n (%) \*** |  |
| Male | 151 (65.4) |
| Female | 80 (34.6) |
| **Age (years) - median [Q1 - Q3]** | 66.0 [58.5 – 65.2] |
| **Type of stroke - n (%)** |  |
| Ischemic | 160 (68.1) |
| Hemorrhagic | 75 (31.9) |
| **Rehabilitation phase - n (%)** |  |
| [0-3 months] early | 173 (73.6) |
| [3-6 months] late | 62 (26.4) |
| **Period - n (%)** |  |
| [2007 – 2015] | 166 (70.6) |
| [2016-2018] | 69 (29.4) |
| **mFAC scale - n (%) \*\*** |  |
| [0-1] | 163 (71.2) |
| [2-8] | 66 (28.8) |

\* Sex: 4 missing data
\*\* mFAC: 6 missing data

We found a median 6MWT before the program of 0 meters (IQR [0-76]) and a median 6MWT of 62 meters (IQR [0-169]) after the program, corresponding to a median gain of 18 meters (IQR [0-74,5]) (p<0,01) (Figure 1).

Table 2 shows the results of the bivariate and multivariate analyses. Two factors were significantly associated with the 6MWT gain:

- Time since onset: Patients in the early rehabilitation phase significantly improved their walking distance on the 6WMT compared with patients in the late rehabilitation phase (40.0 meters IQR [0-85.0] vs. 0 meters IQR [0-20,3] p < 0,001).
- Severity of stroke: mFAC before rehabilitation was also significantly associated with the evolution of the 6WMT. Initially, ambulatory patients had indeed a better recovery than non-ambulatory patients (55.0 meters IQR [22.0-93.0] vs. 0 meters IQR [0-50,0] p < 0,001).

**Table 2:** Bivariate analysis and Multivariate quantile regression analysis of the gain at 6MWT.

| Variable | Bivariate analysis |  | Multivariate quantile regression analysis |
| --- | --- | --- | --- |
|  | Median Gain<br>6MWT [Q1-Q3] | p-value | Coefficient (95% CI) |
| **Sex** |  |  |  |
| Male | 24.0 [0 – 68.0] | 0.416 | 3.22 [- 11.86 ; 19.48] |
| Female* | 11.5 [0 – 78.8] |  |  |
| **Age (years)** | rho** = -0.15 | 0.064 | -0.56 [-0.99 ; 0.16]*** |
| **Type of stroke** |  |  |  |
| Ischemic* | 20.0 [0 – 75.3] | 0.833 | -1.67 [-17.03 ; 10.59] |
| Hemorrhagic | 15.0 [0 – 60.0] |  |  |
| **Rehabilitation phase** |  |  |  |
| [0-3 months] early* | 40.0 [0 – 85.0] | < 0.001 | -21.22 [-44.93 ; -1.86] |
| [3-6 months] late | 0 [0 – 20.3] |  |  |
| **Period** |  |  |  |
| [2007 – 2015] * | 13.0 [0 – 71.5] | 0.271 | 6.22 [-7.99 ; 32.78] |
| [2016-2018] | 43.0 [0 – 80.0] |  |  |
| **mFAC scale** |  |  |  |
| [0-1] * | 0 [0 – 50.0] | < 0.001 | 34.22 [16.20 ; 74.69] |
| [2-8] | 55.0 [22.0 – 93.0] |  |  |

\* Value of Reference
\*\* Correlation coefficient of Spearman
\*\*\* meters per year increase
6MWT : 6 minutes walk test
CI : Confidence Interval
IQR : Interquartile Range; Q1-Q3 first and third quartiles

There is no significant difference in median gain between men and women (24.0 meters IQR [0-68.0] vs. 11,5 meters IQR [0-78.8]; p = 0.4); between ischemic and hemorrhagic strokes (20.0 meters IQR [0-75.3] vs. 15.0 meters IQR [0-60.0]; p = 0.8); and between the pre-2016 and post-2016 periods (14.0 meters IQR [0-71,5] vs. 43.

No significant difference was observed according to age; the correlation coefficient of Spearman was −0.15 by years (p = 0.064). This result was close to significant. After adjustment, the association between the evolution of the 6WMT and, respectively, the rehabilitation phase and severity of stroke before rehabilitation was less strong but remained significant. The median gain in 6WMT was indeed greater at 21 meters for patients in the early rehabilitation phase compared to patients in the late rehabilitation phase (21.22 meters (95% CI, 1.86–44.93)). The median gain of the 6MWT for less severe stroke patients (able to ambulate before the program) was 34 meters longer than non-ambulatory patients (34.22 meters (95% CI, 16.20–74.69).

There was a decrease of 0.56 m per year at the 6MWT, and this result was not significant (95% CI −0.99; 0.16).

The other factors were not significant in the multivariate analysis.

Finally, no significant interaction between the mFAC before rehabilitation and the rehabilitation phases was found, meaning that the effect of the rehabilitation phase on the evolution of the 6WMT was not shown to be different between initially walking patients and non-walking patients.

No significant interaction between the period before or after 2016 and the type of stroke was found.

There was no difference in the effect of period on the evolution of the 6MWT between ischemic or hemorrhagic. We inferred that there was no difference in the effect of period on the evolution of 6MWT in ischemic patients.

In Table 3, we compared the group with missing data with the study group. There was significantly more missing data in the group unable to walk at the onset.

**Table 3:** Comparison of missing data from the 6MWT.

| **Variable** | **Missing data**<br>**N= 72** | **Present Data**<br>**N = 163** | **p-value** |
| --- | --- | --- | --- |
| **Sex - n (%)** |  |  |  |
| Male | 46 (30.4) | 105 (69.6) | 0.942 |
| Female | 24 (30.0) | 56 (70.0) |  |
| **Age (years) - median [Q1 - Q3]** | 66.5 [59.0 – 76.3] | 66.0 [57.0 – 75.0] | 0.354 |
| **Type of stroke - n (%)** |  |  |  |
| Ischemic | 54 (33.8) | 106 (66.3) | 0.131 |
| Hemorrhagic | 18 (24.0) | 57 (76.0) |  |
| **Rehabilitation phase - n (%)** |  |  |  |
| [0-3 months] early | 52 (30.0) | 121 (70.0) | 0.747 |
| [3-6 months] late | 20 (32.3) | 42 (67.7) |  |
| **Period - n (%)** |  |  |  |
| [2007 – 2015] | 50 (30.1) | 116 (69.9) | 0.789 |
| [2016-2018] | 22 (31.9) | 47 (68.1) |  |
| **mFAC scale - n (%)** |  |  |  |
| [0-1] | 54 (33.1) | 109 (66.9) | **0.043** |
| [2-8] | 13 (19.7) | 53 (80.3) |  |

IQR : Interquartile range; Q1-Q3 first and third quartiles

## Discussion

The aim of this retrospective, monocentric study of the effects of robotic gait rehabilitation with Lokomat® in combination with conventional therapy was to describe the evolution of the 6MWT in patients with stroke.

There were 163 patients included in our study.

At the beginning, the median 6MWT was 0 meters (IQR [0-76]), and after the rehabilitation, we found a median 6MWT of 62 meters (IQR [0-169]). The median gain was 18 meters (IQR [0–74.5]). The gain was significant (p<0,001) and was greater in patients in the early rehabilitation phase versus in the late rehabilitation phase (21.22 meters (95% CI [1.86–44.93]) and in ambulatory patients versus non-ambulatory patients at the onset (34.22 meters (95% CI [16.20–74.69]). Although we did not find significant differences with the other factors, we did not have any problems with feasibility.

In previous studies, robotic rehabilitation by Lokomat has not shown a greater effect on walking capacity than conventional rehabilitation (12), but it has shown that it could be safely used with no harm to patient outcomes (14). These studies provide an early look at the efficacy of robotic devices but are limited in the number of participants enrolled. In addition, the details of the training programs at Lokomat were not clear.

In our 11-year study, we have included a large number of patients, and each patient had conventional therapy and the same protocol as Lokomat, and their results were comparable.

To our knowledge, no studies have been conducted analyzing the factors associated with a gain in walking capacity after rehabilitation by Lokomat.

The relevance of an improvement in 6MWT of 18 meters depends on the Minimal Clinically Important Difference (MCID) for 6MWT after a stroke. Fulk (26) calculated the MCID of 65 meters as an improvement in the perception of an important change in walking ability estimated by the patient. Although we found an inferior gain, this aim should be interpreted with caution because their population had a mean of 6MWT at 80 +/- 50 meters initially. In our study, 70% of the patients were unable to walk before the rehabilitation and had a median 6MWT of 0 meters IQR [0–76.0]. It was clinically relevant for a patient to be able to gain 18 meters when they could not walk before rehabilitation (for example, by moving around indoors and doing bed-chair transfers) (28).

Nevertheless, more than 25% of patients had reached the MCID of 65 meters after the program because the third interquartile range was 74.5 meters.

The gain was better when we compared the early versus the late rehabilitation phases. These results are consistent with a Cochrane review (10). All these results are also associated with the concept of brain plasticity; the best time for boosting brain plasticity is within 3 months after a stroke (27). The robotic rehabilitation should start as early as possible, particularly in the early stages of stroke recovery. We also found that the gain on the 6MWT was higher when the patient was ambulatory at the onset. In the Cochrane reviews (10), they found that non-ambulatory patients at the start of the study had greater benefits regarding walking speed after rehabilitation with RAGT. These results are not comparable with ours because walking speed was not calculated on a 6MWT (which assesses walking perimeter) but rather on a 5 or 10-meter walk test (which assesses walking speed).

The motor severity of a stroke was measured by the mFAC scale. Usually, the National Institutes of Health Stroke Scale (NIHSS) score is used to determine the severity of a stroke (28). However, this data was not recorded in our rehabilitation medical records.

In the randomized controlled trial Schwartz and All (29), they describe an improvement of FAC and NIHSS scores in subacute stroke patients using RAGT in comparison with conventional therapy. These results suggest that these two scores evolve together.

Regarding age, we found a median loss per year of 0.56 meters. Although this result is almost significant (95% CI [−0.99; 0.16]), we believe that this loss is not clinically relevant.

There were no feasibility problems in each age group.

We did not find a significant difference in Lokomat rehabilitation between males and females. Although it is generally believed that hemorrhagic stroke survivors have better neurological and functional prognoses than ischemic stroke survivors, a recent study shows that the results are discordant (30). Only one study compared the walking performance of hemorrhagic and ischemic stroke patients in RAGT rehabilitation and found significant improvement at 6MWT in both groups, but the gain was not significantly better in any of them (31). We also did not find any difference between ischemic and hemorrhagic stroke patients. There were no feasibility issues for these groups. A thrombectomy has been shown to improve the prognosis for recovery after an ischemic stroke (24). In 2016, in our region, the practice of thrombectomy was generalized and standardized. This meant that patients for whom thrombectomy was indicated benefited from it more generally than before 2016. We sought to study whether this generalization of thrombectomy had an effect. We did not find a difference, but we estimated the practice of thrombectomy in relation to the period and selected only patients admitted to our center for stroke and not patients who received thrombectomy or not, because we did not have the information. Further research is needed in order to evaluate this point. There was more missing data about the 6MWT in the group unable to walk (mFAC 0-1) at the onset. We have 45 missing data points about the 6MWT before the rehabilitation and 56 after, for a total of 72 patients. These patients were not included in the analysis, so it is possible that the median gain was overestimated.

This limitation is related to the retrospective analysis. Only a prospective study could answer this question.

Because of the retrospective nature of our study, we do not know if all the patients who met the criteria for using the Lokomat benefited from it. The assessment was therapist-dependent. As the years went by, therapists could be less apprehensive about using the Lokomat on a larger population. Also, the Lokomat was not always available, and therefore its use was not possible all the time, for example, during periods of maintenance or when the schedule was full. Thus, there may have been a selection bias.

It is important for future research to focus on the identification of patients for whom an exoskeleton would truly benefit. Our study adds to the emerging literature surrounding the use of exoskeletons in stroke rehabilitation. It provides information regarding the factors of the patients who had the best gain, but prospective and randomized research is needed to confirm these first results. Robotic tools are not only used for gait rehabilitation. They also allow for early verticalization of patients thanks to their system of partially or totally body weight support. It also helps to prevent falls (32) and the complications that can result from them (fractures, post-fall syndrome, etc.) (33).

It allows for a lower intensity of exercise training than conventional physiotherapy. It is proven that RAGT is less energy-consuming and cardiorespiratory-stressful than walking without robot assistance (34) and can be used in patients with cardiovascular diseases. In another study, RAGT was shown to have the potential to improve cardiopulmonary fitness (35). In the future, it would be interesting to study different protocols for well-defined populations in order to offer the most appropriate protocol for the patients.

## Conclusion

Our findings suggest that the use of Lokomat associated with conventional physiotherapy should be used for adults after stroke, particularly in ambulatory adults and in the first three months after a stroke. Larger randomized prospective clinical trials are needed to establish the efficacy of Lokomat training for the recovery of walking after stroke, identify patients for whom it will be highly effective, and propose different protocols according to patient characteristics.

## Data Availability

all data referred to in the manuscript are available

